# Assessment of the performance of Lyoplant^®^ Onlay for duraplasty. An observational, multi-center Post Market Clinical Follow-up study

**DOI:** 10.1101/2022.09.08.22279716

**Authors:** Franziska Greifzu, David Breuskin, Anna Prajsnar, Karsten Geletneky, Frank Bode, Jochen Tuettenberg, Joachim Oertel, Axel Stadie

**Affiliations:** Medical Scientific Affairs, Aesculap AG, Am Aesculap-Platz, 78532 Tuttlingen, Germany; Department of Neurosurgery, Saarland University Medical Center, Saarland University Faculty of Medicine, Kirrberger Straße 100, 66421 Homburg/Saar, Germany; Department of Neurosurgery, Hospital Darmstadt, Grafenstraße 9, 64283 Darmstadt, Germany; Department of Neurosurgery, Hospital Idar-Oberstein, Dr.-Ottmar-Kohler-Straße 2, 55743 Idar-Oberstein, Germany

**Keywords:** Cerebrospinal fluid leak, Collagen implant, Dural substitute, Duraplasty, Lyoplant Onlay, Prospective study, Xenograft

## Abstract

**Objective:** Duraplasty with a dura mater substitute can become necessary after cranial or spinal surgery. Several dura grafts from different materials are available. The goal of this study was to assess the clinical performance of a suturable onlay dura substitute composed of bovine collagen.

**Methods:** A prospective, observational, multicenter, single-arm clinical study with the dura substitute Lyoplant^®^ Onlay was performed. Different performance parameters for safety and performance were assessed. Additionally, postoperative MRI / CT images were evaluated.

**Results:** Data of 61 patients recruited by three study centers were included. Lyoplant^®^ Onlay was used by a variety of surgeons with different levels of experience. Cranial and spinal cases with a wide range of dura defect sizes were included. No reoperation due to a cerebrospinal fluid leakage occurred until discharge (primary endpoint), as well as to the follow-up (about four months postoperatively). The incidence rate of cerebrospinal fluid leakages was 6.6%. All of these adverse events were reported as non-serious complications without the need of a surgical intervention and were resolved without sequelae. The handling properties of Lyoplant^®^ Onlay were positively rated. In external radiologic analysis of postoperative MRI / CT images at discharge and follow-up, additional fluid collection, edema, and swelling were observed. However, they were clinically inapparent and considered normal postsurgical findings.

**Conclusions:** The incidence rate of cerebrospinal fluid leakages was 6.6% and ranged in the values reported in the literature.

These results show that Lyoplant^®^ Onlay is a safe and efficient dura substitute.

**Highlights:**

- Lyoplant^®^ Onlay is a safe and effective bovine dura substitute
- Universal product for onlay or suturable application, cranial and spinal surgeries
- No re-operations because of CSF leakage
- Very good handling characteristics
- Prospective study with 61 patients, 3 study centers and 4 months follow-up

## Introduction

Duraplasty in neurosurgery is used to repair the dura mater in order to avoid complications like cerebrospinal fluid (CSF) leakage. An ideal dura graft should therefore include the following properties: prevention of CSF leakage, biological compatibility, inertness, nonadherence to surrounding tissues, easy handling, cost efficiency, and availability. Different graft materials are available, classified as autograft, allograft, xenograft, and synthetic graft ^1^.

The use of autografts is common, but requires additional preparation for harvesting, including at times a second incision, and may therefore increase the operation time and the morbidity risk for the patient. Additionally, the autograft is usually only available in a limited size. Larger dura defects may therefore not be sufficiently closed. Apart from autografts and allografts, plenty dural substitutes are available, including xenografts mainly produced from equine ^2^, porcine ^3^, or bovine ^4,5^ tissues. Collagen-based products are available in suturable and in onlay versions. Used as an onlay version, they may reduce the operation time ^6^. The application of xenografts is widespread and well-established ^7^. The following problems have been discussed after the use of xenografts: premature absorption of the product, immune reaction against the graft, and nonwatertight CSF closure ^1^.

Lyoplant^®^ Onlay is an absorbable and porous xenograft of bovine pericardium. It can be applied as an onlay device; fixation with sutures is also possible. In an animal study Lyoplant^®^ Onlay showed better handling characteristics and a shorter duraplasty time compared to periosteum, with a trend for better adhesion to dura and CSF tightness compared to another collagen dural graft ^5^.

Here, the safety of Lyoplant^®^ Onlay, was assessed by the incidence of reoperations because of CSF leakage until discharge as a primary variable. Different safety and performance parameters served as secondary variables. To our knowledge, this is the first study to assess Lyoplant^®^ Onlay’s clinical safety and performance.

## Materials and Methods

### Study setting and design

The LYON study (Assessment of the performance of LYoplant^®^ ONlay for Duraplasty) was designed as a prospective, observational, multicenter, single-arm cohort study.

It was performed to assess the safety and performance of Lyoplant^®^ Onlay (Aesculap AG, Tuttlingen, Germany) under routine conditions. This dura substitute is a two-layered implant made of purified collagen from bovine pericardium and bovine split hide and has been available since 2013 (CE-mark, FDA approval). The two layers are not chemically crosslinked. Lyoplant^®^ Onlay can be used as an onlay (with additional sutures if wanted) or sutured in place. It is degraded after implantation. The raw material for Lyoplant^®^ Onlay is imported from New Zealand because bovine pericardium from this country is considered safe with regard to BSE (bovine spongiform encephalopathy). In addition, Lyoplant^®^ Onlay is treated with NaOH during processing to further reduce the theoretical risk of transmission.

The study was presented to the Ethics Committee responsible for each participating center and approved accordingly. Ethics committees that approved the study and the approval number: Ethics Committee Ärztekammer des Saarlandes, Saarbrücken, Germany, Bu 257/15; Ethics Committee Landesärztekammer Rheinland-Pfalz, Mainz, Germany, 837.043.16 (10362); Ethics Committee Landesärztekammer Hessen, Frankfurt am Main, Germany, MC 34/2016. The Ethics Committees reviewed the study protocol, protocol amendment, and the Patient Information Sheet and Consent Form.

All patients gave prior written informed consent. The patient’s consent form had to be signed before the surgery by the patient and the physician. Patients were recruited continuously. Pseudonymized data were stored and analyzed.

This trial was registered prospectively at ClinicalTrials.gov under the registration number NCT02678156 and reported in accordance with the STROCSS and CONSORT guideline, where applicable ^8,9^. Moreover, it followed the principles outlined in the Declaration of Helsinki. A clinical study protocol was developed a priori, but not published.

The study was performed in three neurosurgical centers located in Germany (Saarland University Medical Center Homburg, Klinikum Idar-Oberstein, Klinikum Darmstadt) where Lyoplant^®^ Onlay is used routinely. A fourth center was planned but not initiated since the recruitment was already advanced.

Inclusion and exclusion criteria are given in Table 1.

**Table 1:** Inclusion and exclusion criteria.

|  |  |
| --- | --- |
| **Inclusion criteria** | - Patients undergoing elective cranial or spinal surgery with the need of a duraplasty using Lyoplant Onlay<br>- Written informed consent<br>- Life expectancy > 6 months<br>- Age ≥ 18 years |
| **Exclusion criteria** | - Active local or systemic infections<br>- Open cranial trauma<br>- Open spina bifida<br>- Known hypersensitivity to proteins of bovine origin<br>- Representation by a legal guardian or under involuntary commitment<br>- Pregnancy<br>- Participation in another clinical study<br>- Known primary immunodeficiency |

An interim analysis was performed after about half of the patients completed the study in order to review the data primarily regarding complications.

### Primary and secondary objectives / variables

The safety of Lyoplant^®^ Onlay was assessed by the incidence of reoperations because of CSF leakage until discharge, as a primary variable.

Further safety and performance parameters were used as secondary variables, e.g. incidence of reoperation because of CSF leakage until follow-up, complications, and handling. Routine postoperative MRI (Magnetic Resonance Imaging) / CT (Computed Tomography) images at discharge and follow-up were assessed by an external neuroradiologist for fluid collection, surgical area reaction / edema, and soft tissue swelling. The flow chart of the visits with primary and secondary endpoint is shown in Fig. 1.

**Figure 1.**
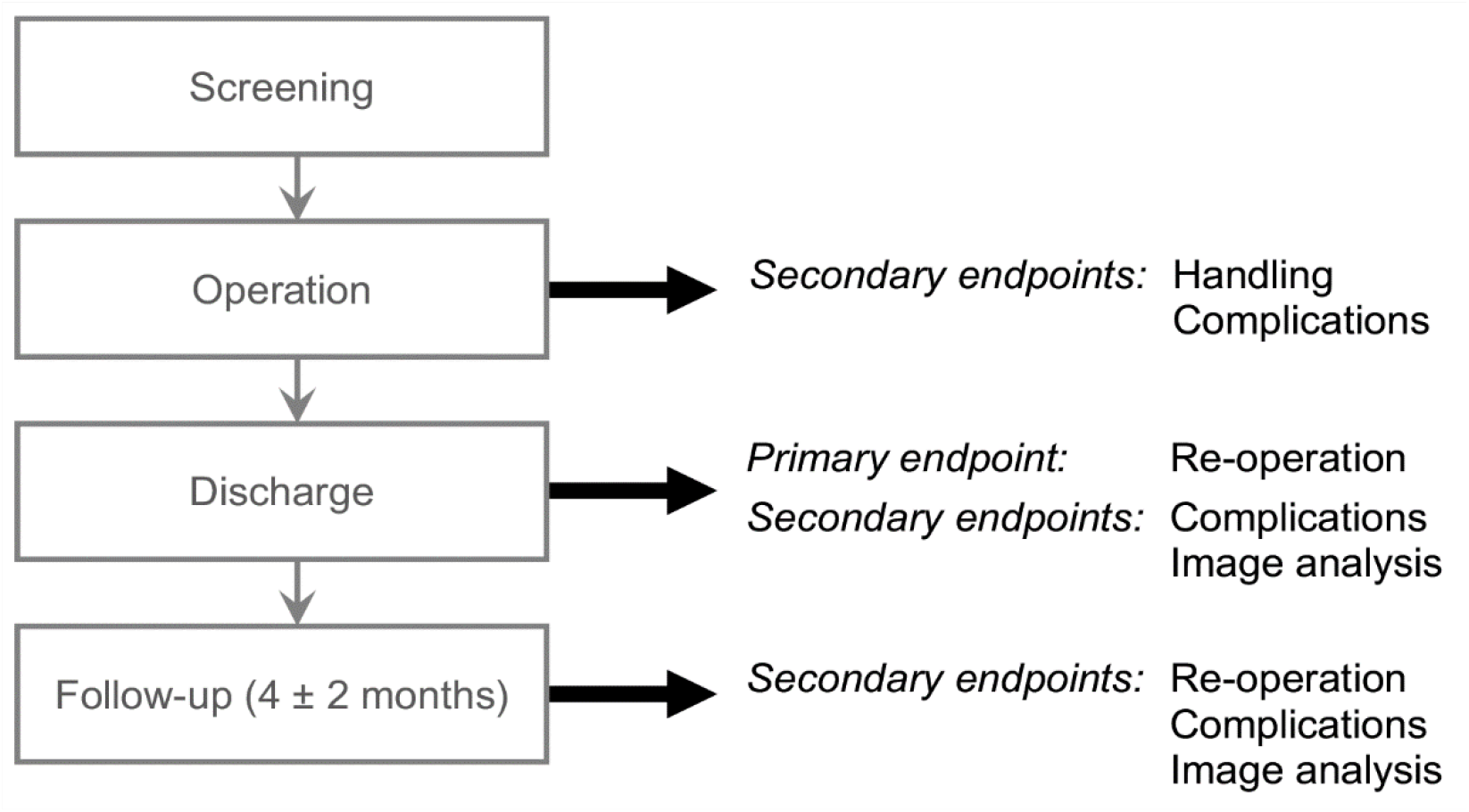
Flow chart of the visits.

### Statistical methods

The sample size of n=60 was anticipated. The average incidence of reoperation due to CSF leakage, 1.9%, was calculated based on publications which reported cranial as well as spinal interventions ^3,10,11^. If there were four or more cases of CSF leakage-related reoperations, then their incidence would become significantly higher than reported (in terms of two-sided 95% Agresti–Coull confidence interval). Thus, four or more events would be critical for the safety of Lyoplant^®^ Onlay.

Statistical analysis was performed by a biometrician with SAS software version 9.4 (SAS Institute, Cary, NC, USA). A multivariable logistic regression model was designated to evaluate interactions of the adverse device effects occurrence with a list of prespecified potential covariates.

## Results

The first patient was enrolled on April 11, 2016; the last completed on February 19, 2018. The recruitment was ended because the planned sample size was reached.

The CONSORT diagram in Fig. 2 shows the number of patients at the different stages. The first study center included 48 patients, the second study center one patient, and the third study center 12 patients. An initial limitation of 30 patients per study center was abrogated to enable sufficient recruitment.

**Figure 2.**
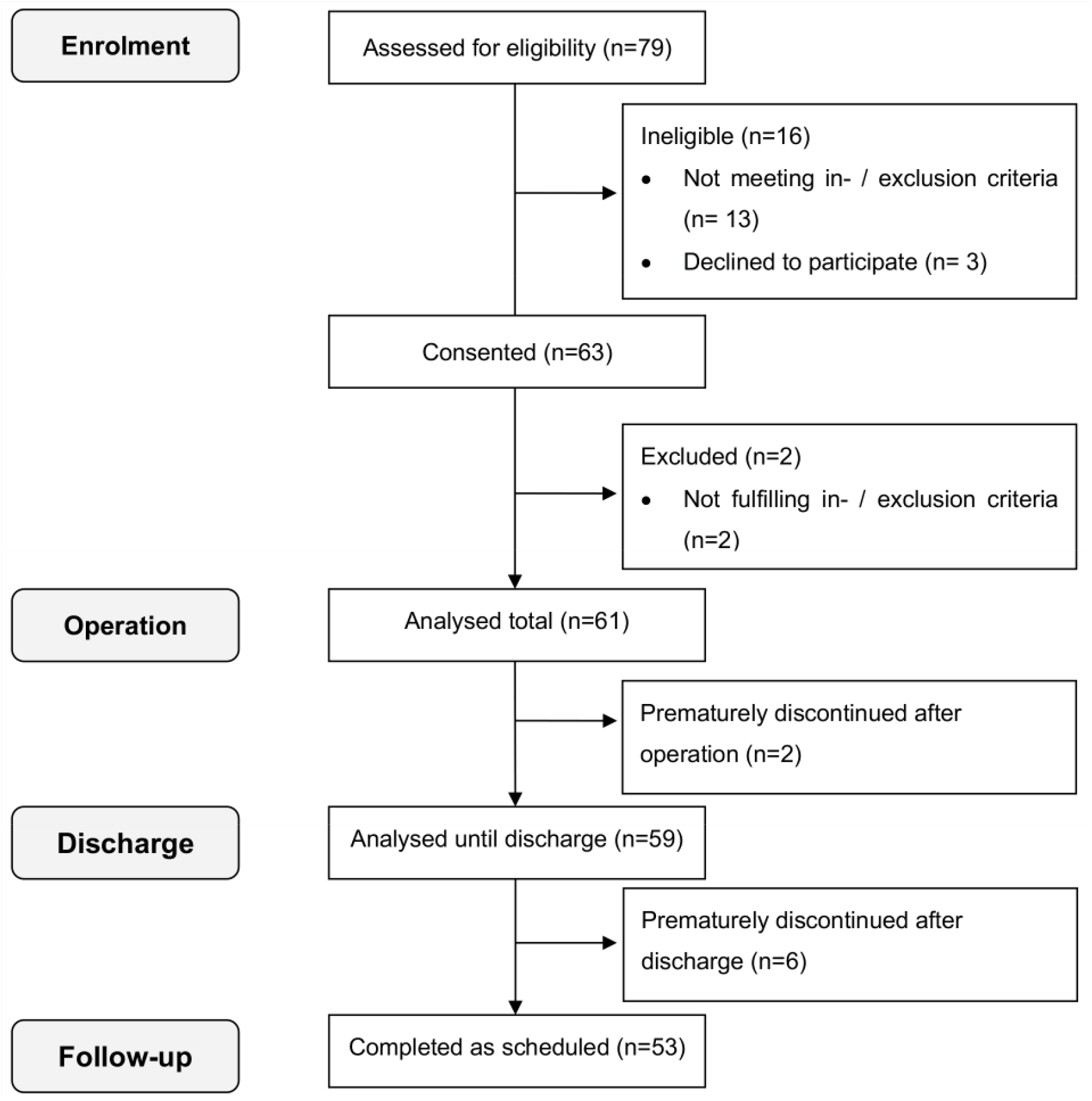
CONSORT flow diagram.

Two patients prematurely discontinued the study after the operation (visit two) and six patients after discharge (visit three). Hence, 59 patients were analyzed until discharge, and 53 patients until the follow-up visit, which completed the study as scheduled including the four ± two months follow-up visit.

Reasons for premature termination were as follows:

Following the surgery visit, one patient had a postoperative rebleeding / edema leading to removal of the Lyoplant^®^ Onlay one day after surgery. This complication had no causal relationship to the product. One patient died after surgery, also not related to the device under investigation.

After discharge, four patients were lost to follow-up. One patient prematurely terminated after discharge due to a wound infection, leading to explantation of the Lyoplant^®^ Onlay. This complication had no causal relationship to the product. During reoperation, it was observed that Lyoplant^®^ Onlay was still intact and in place. Another patient prematurely terminated after discharge for personal reasons.

### Preoperative and intraoperative assessment

Patient (demographic) data are given in Tables 2 and 3.

**Table 2:** Demographic data of the patients. Part I.

|  | Min | Max | Mean | StdDev |
| --- | --- | --- | --- | --- |
| Age [years] | 30 | 84 | 61 | 15 |
| Body Mass Index [kg/m<sup>2</sup>] | 17 | 47 | 29 | 6 |

**Table 3:**
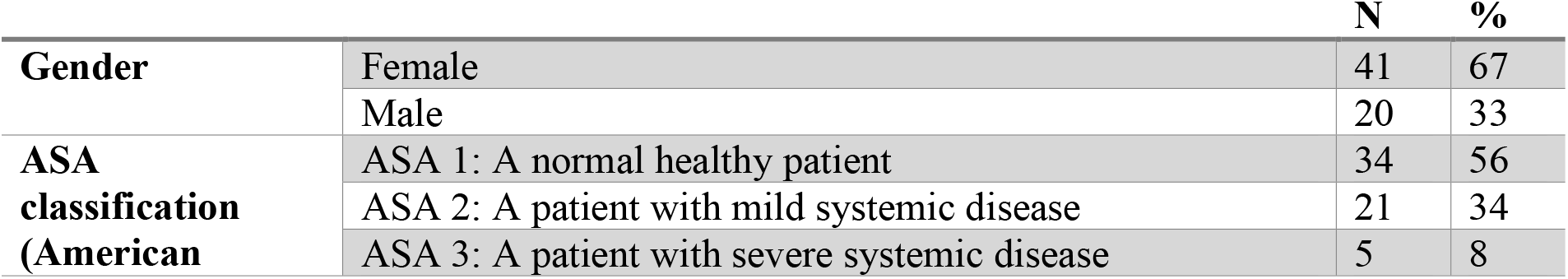
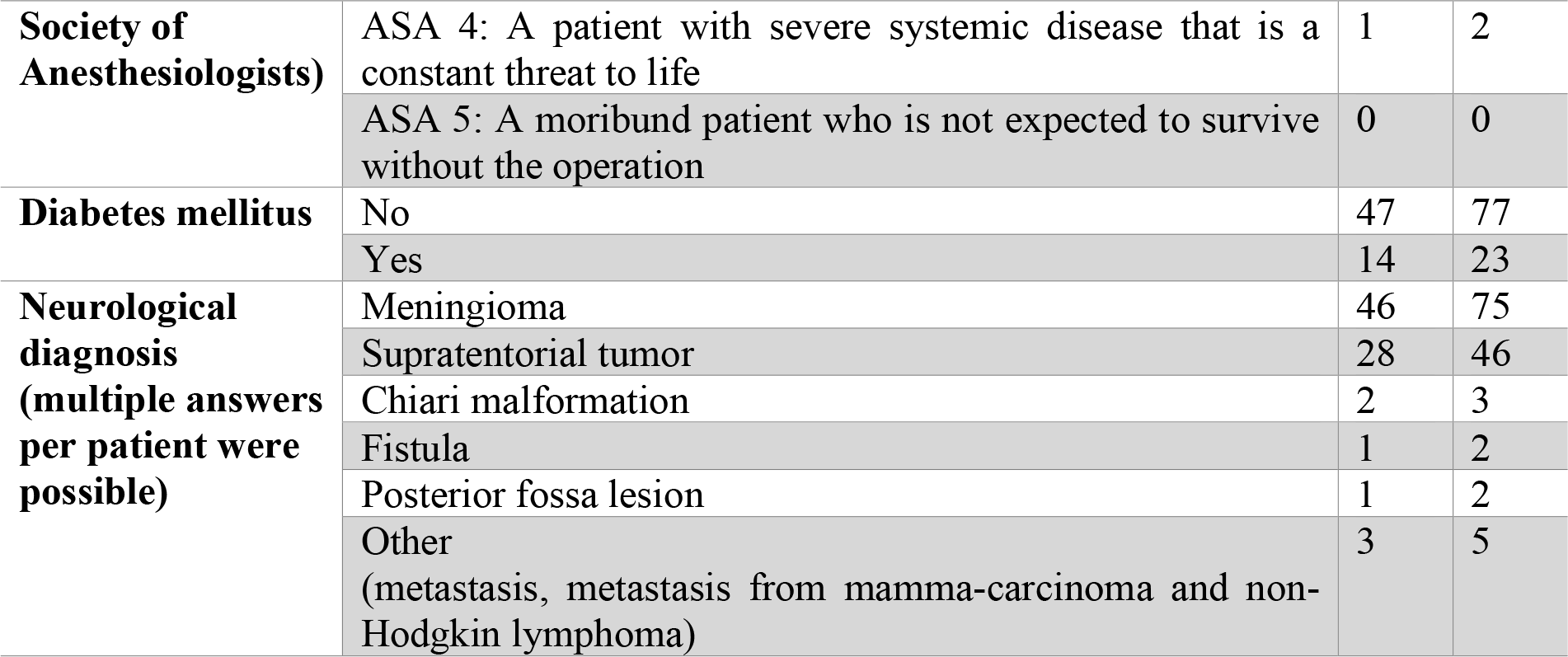
Demographic data of the patients. Part II.

The data in Tables 4 and 5 show that several surgeons with a broad range of experience levels participated in this study.

**Table 4:** Number of surgeons per study site that performed the surgeries within the study.

| **Study site** | **Number of surgeons** |
| --- | --- |
| **001** | 12 surgeons |
| **002** | 1 surgeon |
| **003** | 6 surgeons |
| **Total** | 19 surgeons |

**Table 5:** Surgical experience of the operating surgeons at the start of the study.

| **Surgeons experience** | **Number of surgeons** |
| --- | --- |
| **1 - 2 years** | 1 surgeon |
| **3 - 4 years** | 4 surgeons |
| **5 - 6 years** | 5 surgeons |
| **7 - 10 years** | 2 surgeons |
| **10 - 20 years** | 5 surgeons |
| **> 30 years** | 2 surgeons |
| **Surgeons experience in the application of Lyoplant Onlay** |  |
| **0 times** | 5 surgeons |
| **1 - 5 times** | 8 surgeons |
| **6 - 10 times** | 0 surgeons |
| **> 10 times** | 6 surgeons |

In most cases Lyoplant^®^ Onlay (55 patients) was applied cranially, particularly in the fronto-temporal and in the parietal-occipital or parietal area. In three patients it was applied in the posterior fossa. In three cases Lyoplant^®^ Onlay was applied in the spinal-cervical area. In two of these spinal-cervical cases, the diagnosis was a meningioma, and in one it was Chiari malformation.

The size of the implanted Lyoplant^®^ Onlay after cutting ranged from 3.8 cm^2^ (1.5 × 2.5 cm) to 250.0 cm^2^ (10 × 25 cm) (42.2 ± 35.7 cm^2^). The maximum size resulted from two of the largest Lyoplant^®^ Onlay patches (two times 10.0 × 12.5 cm) sutured together because of the size of the specific dura defect. This was the only case where this combination of two patches was observed. No adverse events related to Lyoplant^®^ Onlay occurred in that patient. However, the implant was removed postoperatively due to a wound infection. Until then Lyoplant^®^ Onlay was still intact and in place.

The size of the dura defect ranged between 0.6 cm^2^ (0.2 × 2.8 cm) and 114.8 cm^2^ (8.5 × 13.5 cm; mean: 22.6 ± 19.8 cm^2^). Lyoplant^®^ Onlay was preferably used in an underlay fashion (application on the dura mater from underneath; 38 patients). Of these, 37 were cranial, out of which one was in the posterior fossa, and one was in the spinal area. The onlay application was used in 21 patients. Of these, 20 applications were in the cranial area (out of which one in the posterior fossa) and one in the spinal area. In two cases (one posterior fossa, one spinal) Lyoplant^®^ Onlay was used onlay as well as underlay (“sandwich-technique”).

Lyoplant^®^ Onlay was additionally fixed in 36 patients with different methods: suture (n=26), bone flap (n=7), fibrin glue (n=3) and other measures (TachoSil, DuraSeal, Spongostan, TachoSil plus Galea; n=14). In 12 patients a combination of methods was used.

In the majority of patients, no drainage was applied (56 patients). Out of the five patients that received a drainage, one had a lumbar drainage, two had a subcutaneous drainage, and two had a subgaleal drainage.

All intraoperative handling parameters were primarily rated as “excellent” or “very good” by the surgeons (Fig. 3) and did not correlate with the experience of the surgeon, neither with the surgical experience in general, nor with the experience in application of Lyoplant^®^ Onlay (p > .05 for all comparisons, Spearman correlation).

**Figure 3.**
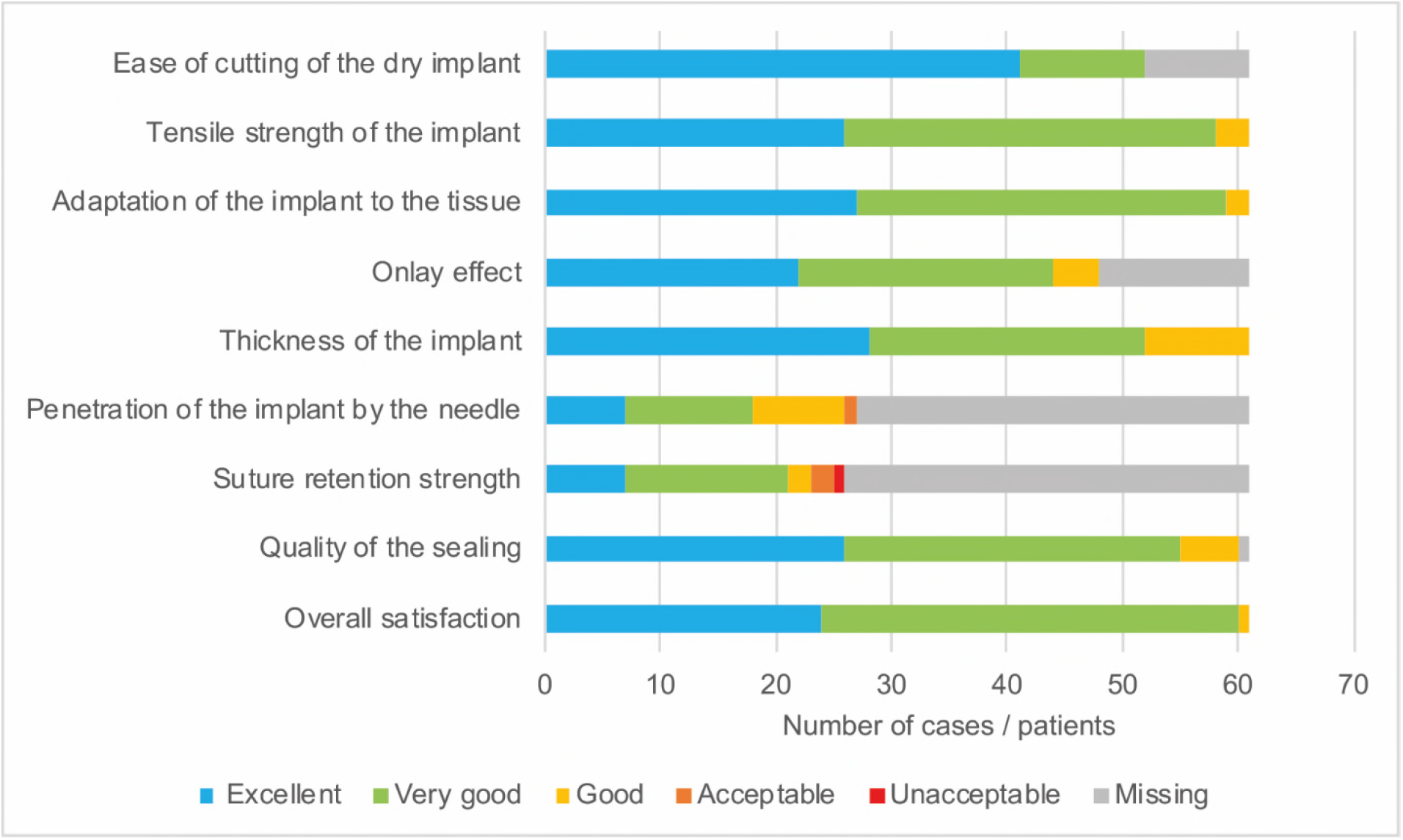
Evaluation of handling characteristics. Evaluation of the intraoperative handling characteristics of Lyoplant^®^ Onlay rated on a five-point Likert Scale, from excellent to unacceptable, with the respective number of cases / patients (frequency). The parameter “onlay effect” was described as no slipping of the implant and sufficient adherence of the implant to the dura. Missing means that the parameter was not assessed / not rated. The parameters “penetration of the implant by the needle” and “suture retention strength” were only assessed when the implant was sutured.

### Discharge and follow-up assessment

The discharge visit took place two to 28 days (six ± four days) after the operation. The follow-up visit was performed 54 to 291 days (about 1.8 months to 9.5 months; 103 ± 41 days) postoperatively. Obviously, some of the follow-up visits took place outside of the planned time frame of four ± two months (n=6). However, since the data still contained valuable information, it was kept.

At discharge, the wound was tight and dry in all 59 patients. Subcutaneous swelling occurred in only four patients. Subcutaneous fluid collection occurred in five patients. In the follow-up visit, the wound was still tight and dry in all assessed patients (53 patients). Subcutaneous swelling occurred in one patient, and subcutaneous fluid collection occurred in two patients.

### Safety

As a primary variable and safety outcome, the incidence of reoperation because of CSF leakage until discharge was evaluated. In none of the patients was the endpoint achieved, neither until discharge nor until the follow-up visit (0.0%).

Table 6 shows the observed complications and their frequency. The majority of complications had no association to Lyoplant^®^ Onlay. Only fluid collection because of CSF leakage was rated as causally related to the device (n = four patients). The surgical area was spinal (n=2) and cranial (n = two). The complications occurred three to 96 days after surgery. In three patients the complication occurred three to seven days after surgery. In the fourth patient it was documented after 95 days (during the follow-up visit). This patient was reoperated earlier (15 days after surgery), due to a postoperative wound infection and meningitis. During that surgery no CSF leakage was observed, and the Lyoplant^®^ Onlay was not removed. In two of these patients no therapeutic measures had to be taken. In the other two patients, nonoperative measures were taken, i.e. lumbar drainage and MRI control after three months. Subsequently, Lyoplant^®^ Onlay related complications were resolved without sequelae.

**Table 6:**
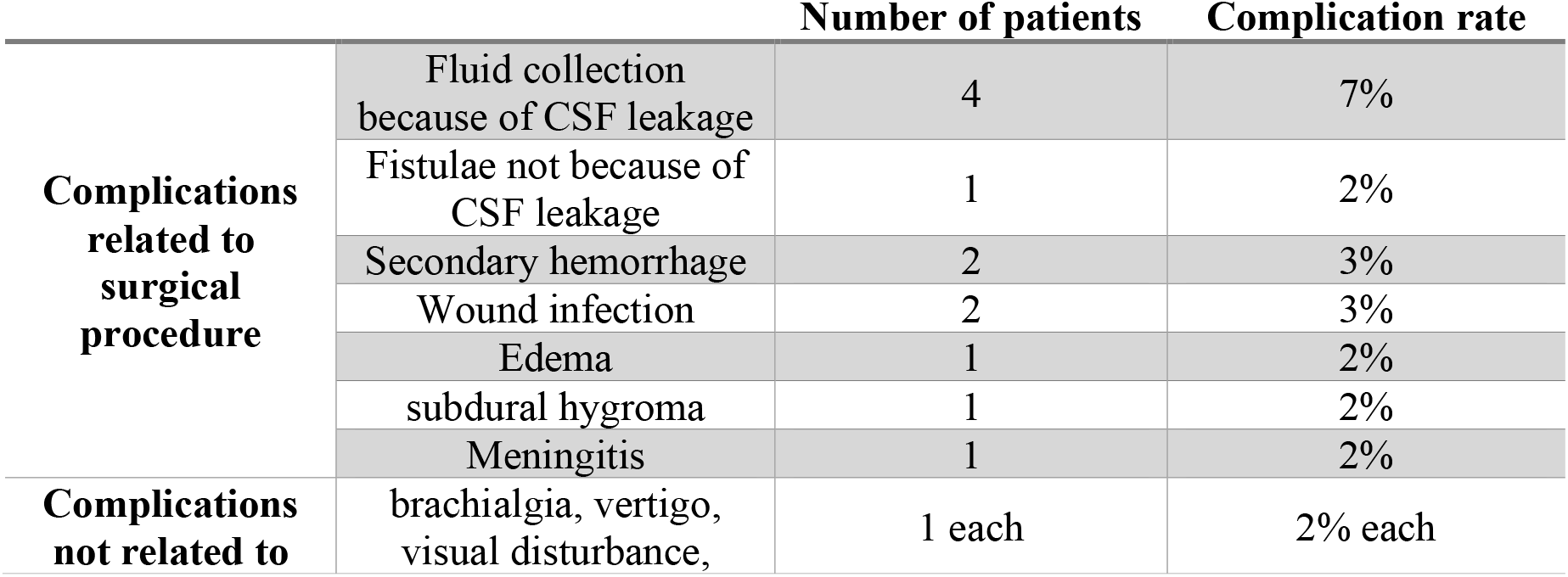
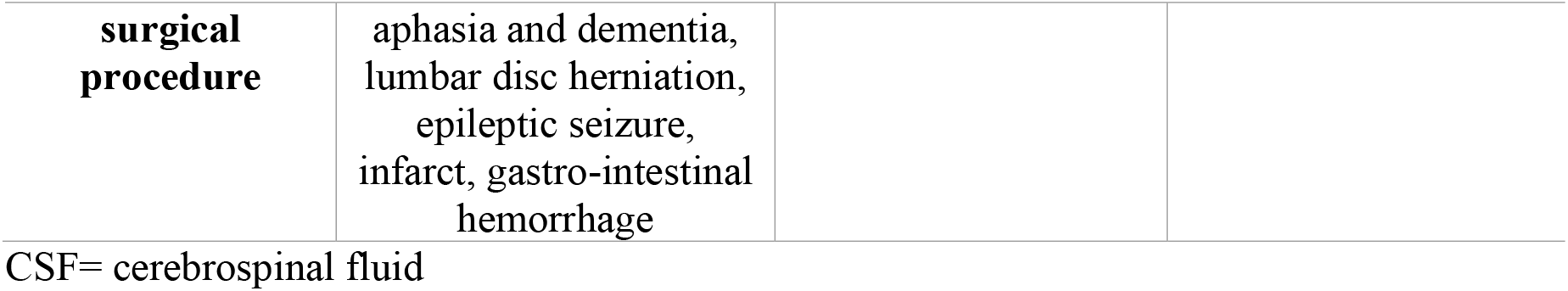
Complications with and without relationship to the surgical procedure (as assessed by the physicians) and the corresponding number of patients and complication rate. Please note that multiple complications per patient were possible.

### Interaction analysis

The analysis of interactions with the primary endpoint (reoperation due to CSF leakage until discharge) became obsolete due to lack of primary events.

A logistic regression model did not show a significant interaction of complications related to Lyoplant^®^ Onlay to any of the tested parameters (p > .05, Table 7).

**Table 7:** The logistic regression model did not show any interaction between the adverse device effect occurrence and tested parameters.

| **Effect** | **p for interaction** |
| --- | --- |
| **Age [years]** | 0.868 |
| **Body Mass Index** | 0.796 |
| **Gender** | 0.980 |
| **Has Lyoplant Onlay been fixed?** | 0.753 |
| **Application onlay** | 0.899 |
| **Application underlay** | 0.905 |
| **Dura defect area (width x length) [cm<sup>2</sup>]** | 0.645 |
| **Surgeons experience [years]** | 0.984 |
| **Surgical area** | 0.453 |
| **Center number** | 0.984 |
| **Rehydration time [seconds]** | 0.936 |
| **Overlap area [cm<sup>2</sup>]** | 0.872 |

### Image analysis

MRI / CT images for the discharge visit were obtained from 47 patients (all CT scan), for the follow-up visit of 44 patients (42x MRI, 2x CT). One study site provided no images. Images were analyzed for fluid collection, edema, and swelling, which were found in several cases. (Results are given in Table 8). Most of them were clinically inapparent and considered normal postsurgical findings by the independent neuroradiologist. Other than CSF accumulations, the observed fluid collections were most likely blood, wound secretion, other liquids, and mistaken residues of hemostyptic products. An unambiguous differentiation was not always possible. Therefore, in most cases, the observed fluid collections are most likely due to other reasons than CSF leakage.

**Table 8:** Results of the analysis of the MRI (Magnetic Resonance Imaging) / CT (Computed Tomography) images at discharge and follow-up.

|  |  | **Discharge** | **Follow-up** |
| --- | --- | --- | --- |
| **Diameter of fluid collection** | < 0.5 cm | 1 patient | 14 patients |
|  | 0.5 – 1.5 cm | 3 patients | 2 patients |
|  | > 1.5 cm | 1 patient | 3 patients |
| **Thickness of surgical area reaction / edema** | < 10 mm | 17 patients | 0 patients |
|  | 10-20 mm | 9 patients | 2 patients |
|  | > 20mm | 3 patients | 0 patients |
| **Thickness of soft tissue-swelling** | < 10 mm | 31 patients | 11 patients |
|  | 10-20 mm | 2 patients | 1 patient |
|  | > 20mm | 1 patient | 1 patient |

## Discussion

The primary variable “incidence of reoperation because of CSF leakage” occurred in none of the patients.

This incidence rate of 0.0% is compared to the average incidence rate of 1.9% calculated from literature values ^3,10,11^. Consequently, we found no evidence for watertightness problems of the Lyoplant^®^ Onlay.

The incident rate of CSF leakage in this study was 6.6%. Reported CSF leakage rates in the literature vary depending on the surgical location, with higher CSF leakage rates in spinal cases. In a review Brandão and colleagues found CSF leakage rates between 0% and 15% following duraplasty, using various dural substitutes ^12^. In their own study they found a CSF leakage rate of 4% for a porcine collagen product and 6% for pericranium or fascia lata for cranial application within a three-month follow-up ^12^. Litvack and colleagues report an overall frequency of CSF leaks of 6.7% in an evaluation of onlay collagen matrix allografts ^13^. The CSF leakage rates within the study ranged from 4.6% up to 11.3% depending on the application site (infratentorial versus supratentorial, significant difference). A clinical study with an onlay collagen graft comparable to Lyoplant^®^ Onlay, reported a CSF leakage rate of 11.9% in posterior fossa surgery^14^. In a prospective case-control study evaluating an onlay collagen matrix for the repair of cranial and spinal dural defects, no CSF leaks occurred ^11^. Another comparable bovine pericardium (collagen) dura graft had a CSF leakage rate of 5.7% in cranial and craniospinal operations ^15^.

In our study, in two of the patients with CSF leakage, Lyoplant^®^ Onlay was implanted in the spine, and in one patient in the posterior fossa region. These areas are more prone to CSF leakage because of higher hydrostatic pressure ^16,17^. Posterior fossa surgery is known to be associated with increased hydrodynamic complication rates ^18,19^. Moskowitz and coworkers described an overall complication rate of 22% and a CSF leakage rate of 11.9% for several types of dural substitutes in posterior fossa surgery ^14^. Walcott and colleagues found a significant relation of the infratentorial operative location to CSF leakage ^20^.

Accordingly, the CSF leakage rate observed in our study is comparable to those described in the literature.

In all cases of CSF leakage, no operative measure had to be taken. They were rated as not serious and resolved without sequelae. Therefore, CSF leakages could be regarded as no immediate patient health risk and are common complications in duraplasty. This findings are in line with a study stating that watertight dural closure is not always necessary, especially in cases of supratentorial, elective surgery ^17^.

No other complications related to Lyoplant^®^ Onlay occurred, especially no foreign body reaction or implant infection developed as described for nonautologous grafts ^21,22^.

Interestingly, the current data revealed that Lyoplant^®^ Onlay was preferably applied in an underlay technique. It is also important to notice that in this form of application the fleece-like porous “dura side” of the implant faces the dura. The underlay use is also described for other (suturable) collagen-derived dura grafts ^13,23^, especially in posterior fossa procedures ^24^. The advantage of this technique is the positive sealing effect of the intracranial pressure, which presses the implant against the dura. Whereas in an onlay application, the pressure rather pushes the graft away from the dura.

In two cases Lyoplant^®^ Onlay was used in a “sandwich-technique”: one patch onlay and one as underlay (one posterior fossa and one spinal). In a clinical study this technique with a combination of autologous dura, dural substitute, and fibrin glue correlated with a significant reduction of CSF leaks in posterior fossa surgery ^25^. Using bovine pericardium, it was also shown as beneficial in terms of blood loss and surgical time during subsequent cranioplasty ^26^. In our study, no interaction between the application technique and CSF leakages were present. However, in certain cases, e.g. spinal or posterior fossa surgery, it might be advantageous to use more elaborate techniques of dural closing.

In about 40% of the patients, Lyoplant^®^ Onlay was not additionally fixed. If fixed, different methods were used. There was no statistical interaction between the fixation and adverse events related to the product. Additional fixation did not increase the watertightness and therefore seems not to be mandatory.

Summarizing, the occurrence of complications seemed to depend on the area of application rather than on the product or its application.

Surgeons with different levels of experience participated in the study, and the occurrence of complications related to Lyoplant^®^ Onlay was not influenced by the surgeon’s level of experience. This suggests that the device can be used with equal safety by experienced as well as by unexperienced surgeons, underlining its ease of use.

The study included also the analysis of routine postoperative MRI / CT images. Most of the observed fluid collection, edema, and swelling were clinically inapparent and considered normal postsurgical findings by the independent neuroradiologist. This has also been described in another study with a dura substitute ^27^.

All intraoperative handling parameters were primarily rated as “excellent” or “very good” and support the product’s suitability as a suturable dura graft, in addition to its onlay / underlay application. Altogether, an “excellent” or “very good” overall handling was reported in 96% of cases. These positive results are noteworthy, since the product was applied by experienced as well as unexperienced surgeons, and also in cases considered more complicated (posterior fossa, spine, for example). According to the previous finding, we conclude that the handling characteristics of Lyoplant^®^ Onlay are very good.

An advantage of onlay grafts is also that they do not require suturing and are therefore time-saving, and consequently, cost-saving ^6,28^. In addition, these grafts can be applied in more difficult surgical locations.

In general, the application method of the dura substitutes is one criteria to differentiate between several products that are available in the market for dura substitution. Lyoplant^®^ Onlay is a dura substitute combining both application methods in one single product. This allows an intraoperative flexibility for the surgeon and makes a universal application possible. Most other products in the market with an onlay application can only be sutured tensionless. This limits the intraoperative flexibility of surgeons because it is recommended to use another product in case an increased tension on the implant is expected.

The possibility for an onlay and / or sutured application in combination with the approved use for cranial and spinal cases enables a familiar use of Lyoplant^®^ Onlay. There is no need for the surgeon to distinguish between different products and their corresponding handling characteristics which makes the intraoperative handling for the whole OR team more consistent. The high tensile strength of Lyoplant^®^ Onlay was evaluated “excellent” or “very good” in 95% of all cases in this clinical trial. This feature supports the prevention of CSF leakage and makes a solid and liquid-tight suturing also under tension possible.

Another advantage of Lyoplant^®^ Onlay is that it is already watertight before implantation due to its compact collagen layers. These collagen layers also integrates with the body’s own connective tissue cells as they act like a scaffold for fibroblast infiltration. In contrast to this, other dura substitutes require a cell infiltration and blood clotting in order to seal the substitute and thus to achieving a watertight closure. For this reason, Lyoplant^®^ Onlay offers the benefit of an immediate watertight dura closure after implantation.

Limitations of our study include limited postoperative follow-up, sample size (especially spinal cases), and number of patients per center.

One study center recruited the majority of patients, because the recruiting rate of this center was higher, and the recruitment started earlier. Therefore, a study site specific bias might be possible. However, within this center 12 different surgeons participated, covering a wide range of surgical experience and experience in the application of Lyoplant^®^ Onlay.

Thus, it can be concluded, that the LYON study presents valid data which can be generalized to different experience levels of surgeons, different application methods, and application sites.

## Conclusion

According to the results of the study, the incidence rate of CSF leakages of the collagen dura substitute Lyoplant^®^ Onlay ranged in the values reported in the literature. No other complications with causal relationship to the product were observed. The handling characteristics are very good. These results show that Lyoplant^®^ Onlay is a safe and efficient dura substitute.

## Supporting information

CONSORT

STROCSS

## Data Availability

All data produced in the present work are contained in the manuscript

## Funding

This work was supported by Aesculap AG, Tuttlingen, Germany

## Role of the funding source

Was involved in study design; in the collection, analysis and interpretation of data; in the writing of the report; and in the decision to submit the article for publication.

## Conflict of interest

Franziska Greifzu is an employee of Aesculap AG.

Axel Stadie has prepared a second clinical study with Aesculap AG. Therefore, he has received expense allowances personally. He is also advising Aesculap AG during the processes of certification of medical products and is therefore receiving expense allowances. Moreover, Axel Stadie has received expense allowances and the traveling costs for presentations and workshops by Aesculap AG.

The other authors have no financial conflict of interest.

## Acknowledgments

The authors would like to thank all the participating surgeons and patients. We also thank Susanne Mathieu, Anna Rasche, and the data management and biometry team at Aesculap AG for their support.

## Notes

### Clinical Trial

NCT02678156

### Funding Statement

This study was funded by Aesculap AG.

### Author Declarations

Ethics Committee/IRB of Aerztekammer des Saarlandes, ethics Committee/IRB of Landesaerztekammer Rheinland-Pfalz and ethics Committee of Landesaerztekammer Hessen gave ethical approval for this work.

## References

1. Sekhar LN, Mai JC. Dural repair after craniotomy and the use of dural substitutes and dural sealants. World Neurosurg. 2013;79(3-4):440–442. doi:10.1016/j.wneu.2011.12.062.

2. Gazzeri R, Neroni M, Alfieri A, et al. Transparent equine collagen biomatrix as dural repair. A prospective clinical study. Acta Neurochir (Wien). 2009;151(5):537–543. doi:10.1007/s00701-009-0290-9.

3. Bejjani GK, Zabramski J. Safety and efficacy of the porcine small intestinal submucosa dural substitute: Results of a prospective multicenter study and literature review. J Neurosurg. 2007;106(6):1028–1033. doi:10.3171/jns.2007.106.6.1028.

4. Filippi R, Schwarz M, Voth D, Reisch R, Grunert P, Perneczky A. Bovine pericardium for duraplasty: Clinical results in 32 patients. Neurosurg Rev. 2001;24(2-3):103–107. doi:10.1007/pl00012392.

5. Neulen A, Gutenberg A, Takács I, et al. Evaluation of efficacy and biocompatibility of a novel semisynthetic collagen matrix as a dural onlay graft in a large animal model. Acta Neurochir (Wien). 2011;153(11):2241–2250. doi:10.1007/s00701-011-1059-5.

6. Danish SF, Samdani A, Hanna A, Storm P, Sutton L. Experience with acellular human dura and bovine collagen matrix for duraplasty after posterior fossa decompression for Chiari malformations. Journal of Neurosurgery: Pediatrics. 2006;104(1):16–20. doi:10.3171/ped.2006.104.1.16.

7. Parízek J, Mĕricka P, Husek Z, et al. Detailed evaluation of 2959 allogeneic and xenogeneic dense connective tissue grafts (fascia lata, pericardium, and dura mater) used in the course of 20 years for duraplasty in neurosurgery. Acta Neurochir (Wien). 1997;139(9):827–838. doi:10.1007/BF01411400.

8. Agha R, Abdall-Razak A, Crossley E, Dowlut N, Iosifidis C, Mathew G. STROCSS 2019 Guideline: Strengthening the reporting of cohort studies in surgery. Int J Surg. 2019;72:156–165. doi:10.1016/j.ijsu.2019.11.002.

9. Schulz KF, Altman DG, Moher D. CONSORT 2010 statement: updated guidelines for reporting parallel group randomised trials. BMJ. 2010;340:c332. doi:10.1136/bmj.c332.

10. Stendel R, Danne M, Fiss I, et al. Efficacy and safety of a collagen matrix for cranial and spinal dural reconstruction using different fixation techniques. J Neurosurg. 2008;109(2):215–221. doi:10.3171/JNS/2008/109/8/0215.

11. Narotam PK, Reddy K, Fewer D, Qiao F, Nathoo N. Collagen matrix duraplasty for cranial and spinal surgery: A clinical and imaging study. J Neurosurg. 2007;106(1):45–51. doi:10.3171/jns.2007.106.1.45.

12. Brandão RACS, Costa BS, Dellaretti MA, Carvalho GTC de, Faria MP, Sousa AA de. Efficacy and safety of a porcine collagen sponge for cranial neurosurgery: a prospective case-control study. World Neurosurg. 2013;79(3-4):544–550. doi:10.1016/j.wneu.2011.08.015.

13. Litvack ZN, West GA, Delashaw JB, Burchiel KJ, Anderson VC. Dural augmentation: part I-evaluation of collagen matrix allografts for dural defect after craniotomy. Neurosurgery. 2009;65(5):890-7; discussion 897. doi:10.1227/01.NEU.0000356970.22315.BC.

14. Moskowitz SI, Liu J, Krishnaney AA. Postoperative complications associated with dural substitutes in suboccipital craniotomies. Neurosurgery. 2009;64(3 Suppl):ons28-33; discussion ons33-4. doi:10.1227/01.NEU.0000334414.79963.59.

15. Anson JA, Marchand EP. Bovine pericardium for dural grafts: clinical results in 35 patients. Neurosurgery. 1996;39(4):764–768. doi:10.1097/00006123-199610000-00025.

16. Rosenwasser RH, Kleiner LI, Krzeminski JP, Buchheit WA. Intracranial pressure monitoring in the posterior fossa: a preliminary report. J Neurosurg. 1989;71(4):503–505. doi:10.3171/jns.1989.71.4.0503.

17. Barth M, Tuettenberg J, Thomé C, Weiss C, Vajkoczy P, Schmiedek P. Watertight dural closure: is it necessary? A prospective randomized trial in patients with supratentorial craniotomies. Neurosurgery. 2008;63(4 Suppl 2):352-8; discussion 358. doi:10.1227/01.NEU.0000310696.52302.99.

18. Djordjevic Z, Milosavlijevic B. Postoperative cerebrospinal fluid fistulas and their treatment. J Neurosurg Sci. 1974;18(2):109–111.

19. Parízek J, Mericka P, Nemecek S, et al. Posterior cranial fossa surgery in 454 children. Comparison of results obtained in pre-CT and CT era and after various types of management of dura mater. Childs Nerv Syst. 1998;14(9):426-38; discussion 439. doi:10.1007/s003810050255.

20. Walcott BP, Neal JB, Sheth SA, et al. The incidence of complications in elective cranial neurosurgery associated with dural closure material. J Neurosurg. 2014;120(1):278–284. doi:10.3171/2013.8.JNS13703.

21. Sun H, Wang H, Diao Y, et al. Large retrospective study of artificial dura substitute in patients with traumatic brain injury undergo decompressive craniectomy. Brain Behav. 2018;8(5):e00907. doi:10.1002/brb3.907.

22. Azzam D, Romiyo P, Nguyen T, et al. Dural Repair in Cranial Surgery Is Associated with Moderate Rates of Complications with Both Autologous and Nonautologous Dural Substitutes. World Neurosurg. 2018;113:244–248. doi:10.1016/j.wneu.2018.01.115.

23. Biroli F, Esposito F, Fusco M, et al. Novel equine collagen-only dural substitute. Neurosurgery. 2008;62(3 Suppl 1):273-4; discussion 274. doi:10.1227/01.neu.0000317404.31336.69.

24. Pettorini BL, Tamburrini G, Massimi L, Paternoster G, Caldarelli M, Di Rocco C. The use of a reconstituted collagen foil dura mater substitute in paediatric neurosurgical procedures--experience in 47 patients. Br J Neurosurg. 2010;24(1):51–54. doi:10.3109/02688690903386991.

25. Heymanns V, Oseni AW, Alyeldien A, et al. Sandwich Wound Closure Reduces the Risk of Cerebrospinal Fluid Leaks in Posterior Fossa Surgery. Clin Pract. 2016;6(2):824. doi:10.4081/cp.2016.824.

26. Nguyen KD, Reddy V, Rahimi SY. Dural Sandwich Technique for Hemicraniectomy and Benefits During Cranioplasty. World Neurosurg. 2019. doi:10.1016/j.wneu.2018.12.162.

27. Messing-Jünger AM, Ibáñez J, Calbucci F, et al. Effectiveness and handling characteristics of a three layer polymer dura substitute: a prospective multicenter. J Neurosurg. 2006;105:701888. doi: 10.3171/jns.2006.105.6.853.

28. Horaczek JA, Zierski J, Graewe A. Collagen matrix in decompressive hemicraniectomy. Neurosurgery. 2008;63(1 Suppl 1):ONS176-81; discussion ONS181. doi:10.1227/01.neu.0000335033.08274.1c.

