## Supplementary material for "Assessment of the performance of Lyoplant^®^ Onlay for duraplasty. An observational, multi-center Post Market Clinical Follow-up study": STROCSS

| The STROCSS 2021 Guideline | | |
|---|---|---|
| **Item no.** | **Item description** | **Page** |
| **TITLE** | | |
| 1 | **Title**<br>• The word cohort or cross-sectional or case-control is included*<br>• Temporal design of study is stated (e.g. retrospective or prospective)<br>• The focus of the research study is mentioned (e.g. population, setting, disease, exposure/intervention, outcome etc.)<br><br>*STROCSS 2021 guidelines apply to cohort studies as well as other observational studies (e.g. cross-sectional, case-control etc.) | Page 1,<br>as applicable |
| **ABSTRACT** | | |
| 2a | **Introduction** – briefly describe:<br>• Background<br>• Scientific rationale for this study<br>• Aims and objectives | 2 |
| 2b | **Methods** - briefly describe:<br>• Type of study design (e.g. cohort, case-control, cross-sectional etc.)<br>• Other key elements of study design (e.g. retro-/prospective, single/multi-centred etc.)<br>• Patient populations and/or groups, including control group, if applicable<br>• Exposure/interventions (e.g. type, operators, recipients, timeframes etc.)<br>• Outcome measures – state primary and secondary outcome(s) | 2 |
| 2c | **Results** - briefly describe:<br>• Summary data with qualitative descriptions and statistical relevance, where appropriate | 2 |
| 2d | **Conclusion** - briefly describe:<br>• Key conclusions<br>• Implications for clinical practice<br>• Need for and direction of future research | 2 |
| **INTRODUCTION** | | |
| 3 | **Introduction** – comprehensively describe:<br>• Relevant background and scientific rationale for study with reference to key literature<br>• Research question and hypotheses, where appropriate<br>• Aims and objectives | 3 |
| **METHODS** | | |
| 4a | **Registration**<br>• In accordance with the Declaration of Helsinki*, state the research registration number and where it was registered, with a hyperlink to the registry entry (this can be obtained from ResearchRegistry.com, ClinicalTrials.gov, ISRCTN etc.)<br>• All retrospective studies should be registered before submission; it should be stated that the research was retrospectively registered<br><br>** “Every research study involving human subjects must be registered in a publicly accessible database before recruitment of the first subject”* | 4 |

| | | |
|---|---|---|
| 4b | **Ethical approval**<br>• Reason(s) why ethical approval was needed<br>• Name of body giving ethical approval and approval number<br>• Where ethical approval wasn't necessary, reason(s) are provided | 4 |
| 4c | **Protocol**<br>• Give details of protocol (*a priori* or otherwise) including how to access it (e.g. web address, protocol registration number etc.)<br>• If published in a journal, cite and provide full reference | (4-6)<br>*protocol was not published* |
| 4d | **Patient and public involvement in research**<br>• Declare any patient and public involvement in research<br>• State the stages of the research process where patients and the public were involved (e.g. patient recruitment, defining research outcomes, dissemination of results etc.) and describe the extent to which they were involved. | N.A.<br>*(no patient or public involvement in research )* |
| 5a | **Study design**<br>• State type of study design used (e.g. cohort, cross-sectional, case-control etc.)<br>• Describe other key elements of study design (e.g. retro-/prospective, single/multi-centred etc.) | 4 |
| 5b | **Setting and timeframe of research** – comprehensively describe:<br>• Geographical location<br>• Nature of institution (e.g. primary/secondary/tertiary care setting, district general hospital/teaching hospital, public/private, low-resource setting etc.)<br>• Dates (e.g. recruitment, exposure, follow-up, data collection etc.) | 4, 7 |
| 5c | **Study groups**<br>• Total number of participants<br>• Number of groups<br>• Detail exposure/intervention allocated to each group<br>• Number of participants in each group | 7-8 |
| 5d | **Subgroup analysis** – comprehensively describe:<br>• Planned subgroup analyses<br>• Methods used to examine subgroups and their interactions | N.A. |
| 6a | **Participants** – comprehensively describe:<br>• Inclusion and exclusion criteria with clear definitions<br>• Sources of recruitment (e.g. physician referral, study website, social media, posters etc.)<br>• Length, frequency and methods of follow-up (e.g. mail, telephone etc.) | 5, Table 1, Figure 1 |
| 6b | **Recruitment** – comprehensively describe:<br>• Methods of recruitment to each patient group (e.g. all at once, in batches, continuously till desired sample size is reached etc.)<br>• Any monetary incentivisation of patients for recruitment and retention should be declared; clarify the nature of any incentives provided<br>• Nature of informed consent (e.g. written, verbal etc.)<br>• Period of recruitment | 4, 7 |
| 6c | **Sample size** – comprehensively describe:<br>• Analysis to determine optimal sample size for study accounting for population/effect size<br>• Power calculations, where appropriate<br>• Margin of error calculation | 6 |
| **METHODS - INTERVENTION AND CONSIDERATIONS** | | |

| | | |
|---|---|---|
| 7a | **Pre-intervention considerations** – comprehensively describe:<br>• Preoperative patient optimisation (e.g. weight loss, smoking cessation, glycaemic control etc.)<br>• Pre-intervention treatment (e.g. medication review, bowel preparation, correcting hypothermia/-volemia/-tension, mitigating bleeding risk, ICU care etc.) | N.A.<br>*(No patient optimisation or specific pre-intervention treatment, but data about health status collected)* |
| 7b | **Intervention** – comprehensively describe:<br>• Type of intervention and reasoning (e.g. pharmacological, surgical, physiotherapy, psychological etc.)<br>• Aim of intervention (preventative/therapeutic)<br>• Concurrent treatments (e.g. antibiotics, analgesia, anti-emetics, VTE prophylaxis etc.)<br>• Manufacturer and model details, where applicable | 4+5<br>*(where applicable)* |
| 7c | **Intra-intervention considerations** – comprehensively describe:<br>• Details pertaining to administration of intervention (e.g. anaesthetic, positioning, location, preparation, equipment needed, devices, sutures, operative techniques, operative time etc.)<br>• Details of pharmacological therapies used, including formulation, dosages, routes, and durations<br>• Figures and other media are used to illustrate | 4-6<br>*(where applicable)* |
| 7d | **Operator details** – comprehensively describe:<br>• Requirement for additional training<br>• Learning curve for technique<br>• Relevant training, specialisation and operator's experience (e.g. average number of the relevant procedures performed annually) | *No training needed. Experience of surgeons and interaction analysis included.*<br>9, 10<br>Table 4 and Table 5 |
| 7e | **Quality control** – comprehensively describe:<br>• Measures taken to reduce inter-operator variability<br>• Measures taken to ensure consistency in other aspects of intervention delivery<br>• Measures taken to ensure quality in intervention delivery | N.A.<br>*(observational study in clinical routine)* |
| 7f | **Post-intervention considerations** – comprehensively describe:<br>• Post-operative instructions (e.g. avoid heavy lifting) and care<br>• Follow-up measures<br>• Future surveillance requirements (e.g. blood tests, imaging etc.) | 5<br>*(partially, no special treatment necessary)* |
| 8 | **Outcomes** – comprehensively describe:<br>• Primary outcomes, including validation, where applicable<br>• Secondary outcomes, where appropriate<br>• Definition of outcomes<br>• If any validated outcome measurement tools are used, give full reference<br>• Follow-up period for outcome assessment, divided by group | 5-6<br>Figure 1 |
| 9 | **Statistics** – comprehensively describe:<br>• Statistical tests and statistical package(s)/software used | 6 |

| | | |
|---|---|---|
| | • Confounders and their control, if known<br>• Analysis approach (e.g. intention to treat/per protocol)<br>• Any sub-group analyses<br>• Level of statistical significance | *(where applicable)* |
| **RESULTS** | | |
| 10a | **Participants** – comprehensively describe:<br>• Flow of participants (recruitment, non-participation, cross-over and withdrawal, with reasons). Use figure to illustrate.<br>• Population demographics (e.g. age, gender, relevant socioeconomic features, prognostic features etc.)<br>• Any significant numerical differences should be highlighted | 7-9<br>Figure 2, Table 2, Table 3 |
| 10b | **Participant comparison**<br>• Include table comparing baseline characteristics of cohort groups<br>• Give differences, with statistical relevance<br>• Describe any group matching, with methods | N.A.<br>*(no comparison, single-arm study)* |
| 10c | **Intervention** – comprehensively describe:<br>• Degree of novelty of intervention<br>• Learning required for interventions<br>• Any changes to interventions, with rationale and diagram, if appropriate | 4-5<br>*(partially, established clinical routine)* |
| 11a | **Outcomes** – comprehensively describe:<br>• Clinician-assessed and patient-reported outcomes for each group<br>• Relevant photographs and imaging are desirable<br>• Any confounding factors and state which ones are adjusted | 10, 13-14<br>*(handling evaluation, MRI / CT images)* |
| 11b | **Tolerance** – comprehensively describe:<br>• Assessment of tolerability of exposure/intervention<br>• Cross-over with explanation<br>• Loss to follow-up (fraction and percentage), with reasons | 7-8<br>Figure 2<br>*(where applicable)* |
| 11c | **Complications** – comprehensively describe:<br>• Adverse events and classify according to Clavien-Dindo classification*<br>• Timing of adverse events<br>• Mitigation for adverse events (e.g. blood transfusion, wound care, revision surgery etc.)<br><br>*Dindo D, Demartines N, Clavien P-A. Classification of Surgical Complications. A New Proposal with Evaluation in a Cohort of 6336 Patients and Results of a Survey. Ann Surg. 2004; 240(2): 205-213 | 12-14<br>Table 6 |
| 12 | **Key results** – comprehensively describe:<br>• Key results with relevant raw data<br>• Statistical analyses with significance<br>• Include table showing research findings and statistical analyses with significance | 7-14 |
| **DISCUSSION** | | |
| 13 | **Discussion** – comprehensively describe:<br>• Conclusions and rationale<br>• Reference to relevant literature<br>• Implications for clinical practice<br>• Comparison to current gold standard of care<br>• Relevant hypothesis generation | 14-18 |

| | | |
|---|---|---|
| 14 | **Strengths and limitations** – comprehensively describe:<br>• Strengths of the study<br>• Weaknesses and limitations of the study and potential impact on results and their interpretation<br>• Assessment and management of bias<br>• Deviations from protocol, with reasons | 17 |
| 15 | **Relevance and implications** – comprehensively describe:<br>• Relevance of findings and potential implications for clinical practice<br>• Need for and direction of future research, with optimal study designs mentioned | 14-18<br>*(except for future research since open question was answered)* |
| **CONCLUSION** | | |
| 16 | **Conclusions**<br>• Summarise key conclusions<br>• Outline key directions for future research | 18 |
| **DECLARATIONS** | | |
| 17a | **Conflicts of interest**<br>• Conflicts of interest, if any, are described | 19 |
| 17b | **Funding**<br>• Sources of funding (e.g. grant details), if any, are clearly stated<br>• Role of funder | 19 |
| 17c | **Contributorship**<br>• Acknowledge patient and public involvement in research; report the extent of involvement of each contributor | N.A.<br>*(no patient and public involvement)* |
